# TAVI PACER: A two-step risk score for prediction of permanent pacemaker implantation after TAVI

**DOI:** 10.1101/2024.08.17.24311901

**Authors:** A. Janiszewski, J. Lueg, D. Schulze, B. Juri, L. Morell, M. Hajduczenia, P. Hennig, A. Erbay, A. Lembcke, S. Niehues, U. Landmesser, K. Stangl, D. Leistner, V. Tscholl, H. Dreger

## Abstract

**Background:** The need for permanent pacemaker implantation (PPMI) remains one of the most frequent complications after transcatheter aortic valve implantation (TAVI). This study aimed to develop a novel, two-step risk score to predict PPMI probability after TAVI and to implement it into a user-friendly website. Our risk score addresses the gap in existing data on current prosthesis generations and provides a new and clinically motivated approach to calculating the risk for PPMI.

**Methods:** Between January 2019 and December 2020, 1039 patients underwent TAVI at our institution. We retrospectively evaluated clinical, electrocardiographic, echocardiographic, computed tomographic, and periprocedural data. Patients with prior PPMI were excluded. We developed a prediction model for the occurrence of PPMI, initially using 55 patient and procedural characteristics.

**Results:** We included 836 patients (mean age 80.2 ± 9.1 years, 50.5% female), among whom 140 patients (16.6%) needed PPMI within 30 days after TAVI. In the first step, the TAVI PACER score calculates an individual risk for PPMI, including 14 preprocedural risk factors such as preexisting right bundle branch block, atrioventricular block, left bundle branch block, bradycardia, interventricular septum thickness in diastole, NYHA class, and aortic annulus perimeter. In the second step, intraprocedural variables are included to demonstrate how PPMI risk can vary based on the chosen valve type and implantation depth. The TAVI PACER score can predict PPMI with a sensitivity of 76% and specificity of 72% (AUC = 0.8).

**Conclusions:** The TAVI PACER score provides a novel tool for daily clinical practice, which predicts the individual risk for PPMI after TAVI based on various patient and two procedural characteristics.

**What Is Known:** High-grade conduction disturbances are among the most common and clinically significant complications following TAVI. While recent research has extensively explored the factors leading to an increased risk of pacemaker implantation after TAVI, an established scoring system that considers individual patient risk profiles and predicts the likelihood of requiring a pacemaker before the procedure is still lacking.

**What the Study Adds:** We developed a new two-step risk score called the TAVI PACER Score, which aims to close the gap in existing data on current prosthesis generations. In the first step, it considers various patient characteristics, and in the second step, it evaluates the impact of different procedural options on the necessity for pacemaker implantation. By calculating the individual pacemaker risk before the procedure and integrating it into treatment planning, our risk score aims to improve transparency in patient care, facilitate shared decision-making, and ultimately enhance the treatment outcomes of TAVI.

## Introduction

Over the past two decades, transcatheter aortic valve implantation (TAVI) has emerged as a safe and effective treatment for patients with severe symptomatic aortic stenosis and high risk for cardiac surgery.^1^ Since its first implementation in 2002, TAVI and its outcomes have steadily improved.^2,3^ Even in patients with low surgical risk, current data show that TAVI notinferior to conventional surgical aortic valve replacement in terms of one-year mortality.^4,5^ However, due to the proximity of the atrioventricular node and the His bundle to the landing zone of the transcatheter heart valve (THV), rhythmological complications remain common with TAVI.^6^ Postoperative permanent pacemaker implantation (PPMI) is required in 3.4% to 25.9% of patients undergoing TAVI, which is significantly higher than after surgical aortic valve replacement.^7,8^ Numerous individual risk constellations contribute to the heart team’s interdisciplinary decision-making process, highlighting the importance of patient-centered precision medicine. Although the factors leading to an increased risk of PPMI have already been examined in several studies, an established risk score to estimate the individual risk of pacemaker implantation after TAVI is still lacking.^9–10^ Two published risk scores^11,12^ have not gained acceptance in everyday clinical practice yet due to their underperformance in external validation cohorts.^13,14^ Recently, Lauten et al. renewed their call for the development of a novel straightforward risk score to predict pacemaker implantation following TAVI.^15^

Based on data from a contemporary TAVI population, our study aimed to develop an innovative tool that assists interventional cardiologists in both selecting the appropriate THV and determining the intraoperative implantation strategy, considering the preexisting individual risk for PPMI. Our risk score is thus intended to facilitate decision-making processes within the heart team and to support informed decision-making between patients and their physicians.

## Materials and Methods

### Study Population

Between January 2019 and December 2020, 1039 patients underwent TAVI at our center. After excluding patients with previous device implantation, valve-in-valve procedure, and patients who had received rarely used or discontinued valves (e.g. ALLEGRA, NVT AG, Morges, Switzerland; LOTUS Edge, Boston Scientific, Natick, Massachusetts; CENTERA, Edwards Lifesciences, Irvine, California), a total of 836 patients were included in the analysis (Figure 1). Following current guidelines, every patient undergoing TAVI received an initial assessment by a multidisciplinary heart team. This study conforms to the principles outlined in the 2014 Declaration of Helsinki. The Ethics Committee at the Charité Mitte Campus of the Charité Ethics Committee (management: Katja Orzechowski, Charitéplatz 1, 10117 Berlin) gave ethical approval for this work on 22^nd^ december 2021 (EA1/341/21).

**Figure 1:**
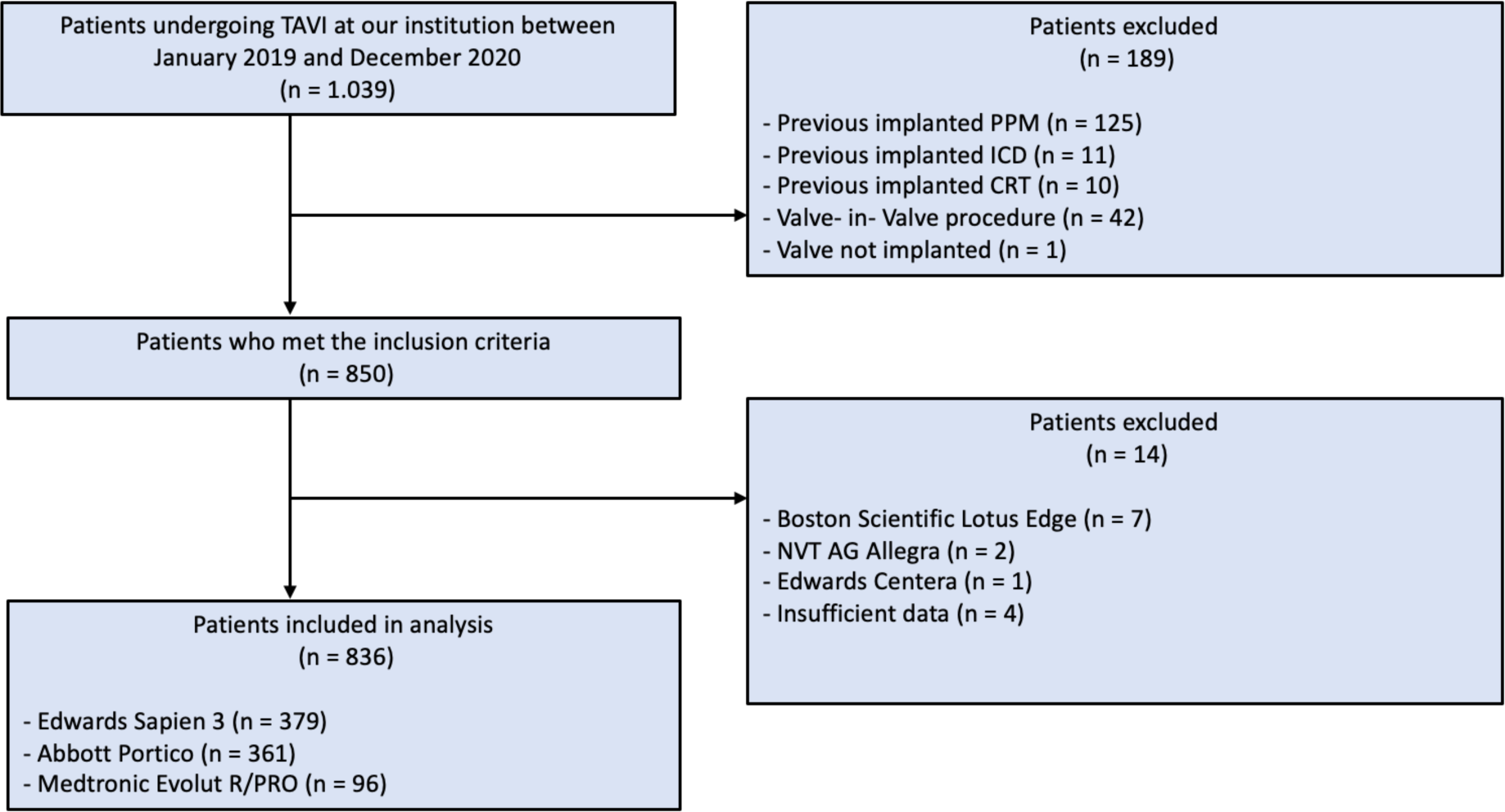
Study workflow.

### Procedures

Patient baseline data, preoperative echocardiographic data, and postoperative complications were retrospectively obtained from electronic medical records. Twelve-lead electrocardiogram (ECG) analysis was performed manually at baseline and within 48 hours after TAVI. Conduction disorders were defined according to the standard definitions of the American Heart Association, the American College of Cardiology Foundation, and the recommendations of the Heart Rhythm Society for the standardization and interpretation of electrocardiograms.^16^ Aortic valve assessment was performed using Pie Medical Imaging’s 3Mensio® software at 25% to 40% of the cardiac cycle. Computed tomography (CT) data was also used to assess THV calcification, to measure the membranous septum length, and to calculate valve oversizing. The implantation depth of the prosthesis in the LVOT was measured using the images of stored intraprocedural fluoroscopic aortography. The measurements were performed with MERLIN Diagnostic software (Phönix-PACS GmbH, Freiburg, Germany). The portion of the prosthetic stent located below the annulus in the left ventricle was measured orthogonally to the aortic annulus at the noncoronary cusp (NCC) and left coronary cusp (LCC). During data aquisition, all investigators were blinded to the postoperative need for a pacemaker. The data was collected retrospectively.

### Statistical analysis

Normally distributed continuous variables are presented as mean ± standard deviation, while non-normally distributed continuous variables are reported as median with interquartile range (IQR). Categorical variables are shown as absolute (n) and relative frequencies (%).

For predicting PPMI, a total of 55 variables were considered. For variable selection, we ran tailored logistic regression models after imputing missing values. Of all data entries, 5.3% were missing. We used multiple imputation with 100 imputed datasets. Lasso regression was used to identify relevant predictors. For each imputation, the recovered relevant predictors under minimal penalty were recorded after running a 10-fold cross-validation. Predictors were selected if they were included in the final model in at least 50% of all imputations. To further improve the final predictive model, all first-order interactions of the preselected variables from the first step were scanned and added to the final model. The predictive quality of the model was assessed by estimating the likelihood of post-TAVI pacemaker implantation using a logistic regression model and calculating the receiver operating characteristic curve for the derived score. The area under the curve (AUC) served as an indicator of model fit (Figure 2). Again, these analyses were pooled for all imputations. All statistical analyses were conducted using R Statistical Software (version 4.2.1; Core Team 2022).

**Figure 2:**
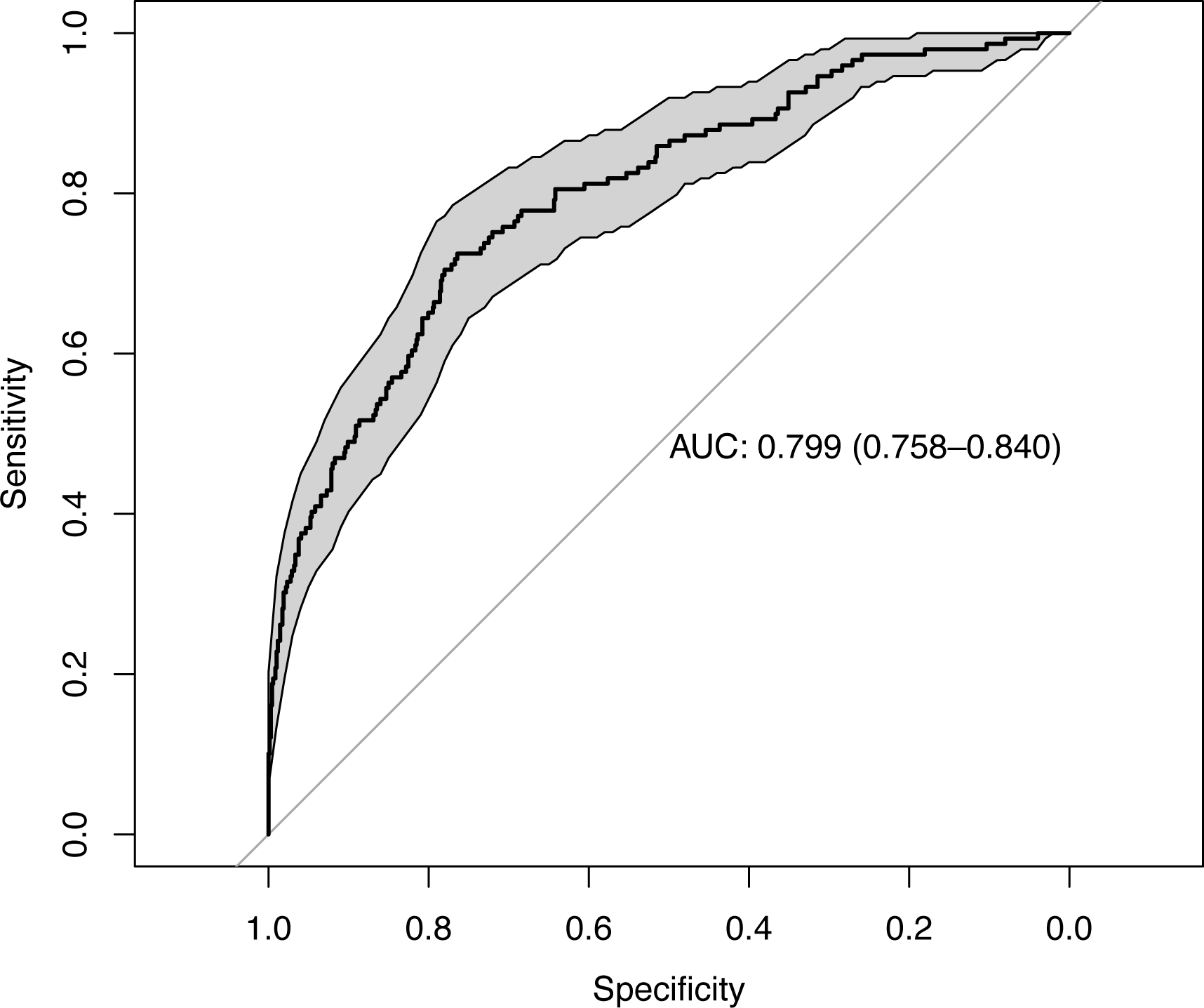
Receiver operating characteristic curve analysis for the final model including interactions.

## Results

### Patient Characteristics

836 patients were included with a mean age of 80.3 ± 9.1 years and a balanced sex ratio (50.6% female, n = 423). The included patients exhibited a low surgical risk [Society of Thoracic Surgeons score 3.4% (IQR 2.3 - 5.2)], along with severe aortic valve stenosis characterized by a mean pressure gradient of 42.0 ± 14.4 mmHg and an aortic valve area of 0.74 ± 0.20 cm^2^. Additional clinical and procedural characteristics are shown in Table 1. Within 30 days after TAVI, 149 patients (17.8%) underwent PPMI, primarily due to third-degree atrioventricular block (AV block) (69%, n = 102). Other indications for PPMI included bradyarrhythmia (14.8%, n = 22), second-degree AV block type Mobitz II (10.7%, n = 16), and sinus node dysfunction (3.4%, n = 5).

**Table 1:**
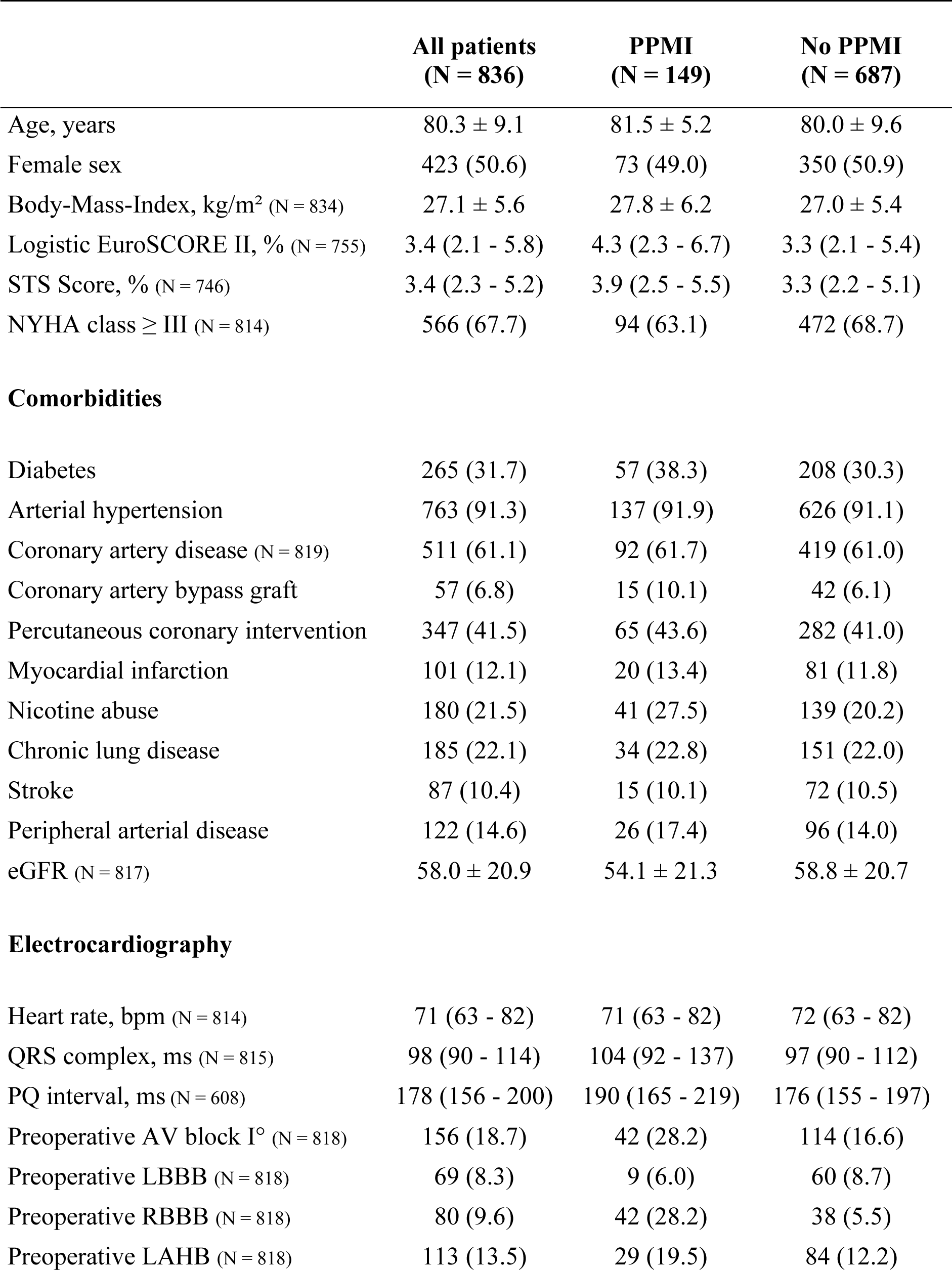
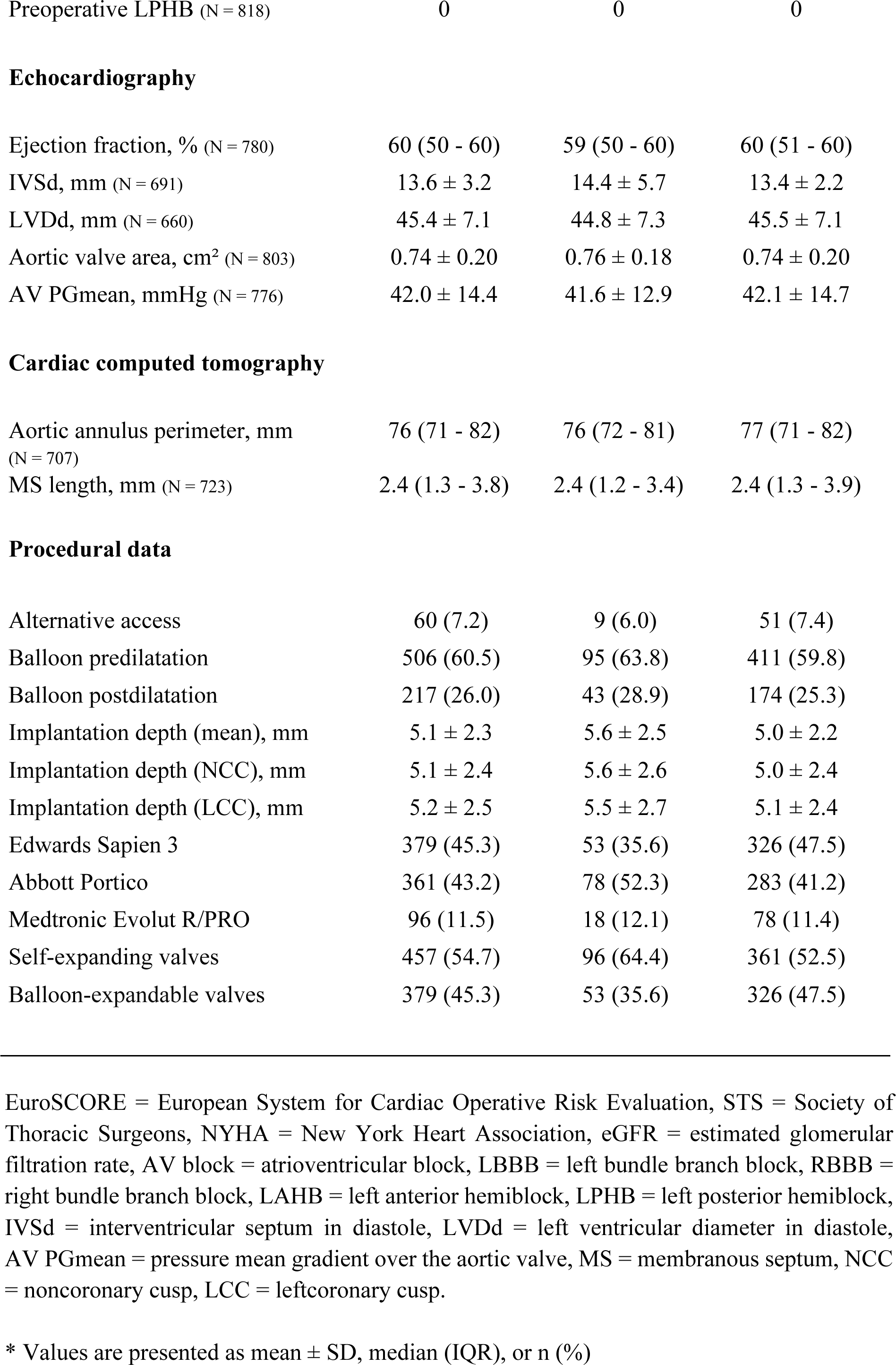
Comparison of Clinical and Procedural Characteristics between patients with and without PPMI after TAVI.

|  | All patients<br>(N = 836) | PPMI<br>(N = 149) | No PPMI<br>(N = 687) |
| --- | --- | --- | --- |
| Age, years | 80.3 ± 9.1 | 81.5 ± 5.2 | 80.0 ± 9.6 |
| Female sex | 423 (50.6) | 73 (49.0) | 350 (50.9) |
| Body-Mass-Index, kg/m<sup>2</sup> (N = 834) | 27.1 ± 5.6 | 27.8 ± 6.2 | 27.0 ± 5.4 |
| Logistic EuroSCORE II, % (N = 755) | 3.4 (2.1 - 5.8) | 4.3 (2.3 - 6.7) | 3.3 (2.1 - 5.4) |
| STS Score, % (N = 746) | 3.4 (2.3 - 5.2) | 3.9 (2.5 - 5.5) | 3.3 (2.2 - 5.1) |
| NYHA class ≥ III (N = 814) | 566 (67.7) | 94 (63.1) | 472 (68.7) |
| Comorbidities |  |  |  |
| Diabetes | 265 (31.7) | 57 (38.3) | 208 (30.3) |
| Arterial hypertension | 763 (91.3) | 137 (91.9) | 626 (91.1) |
| Coronary artery disease (N = 819) | 511 (61.1) | 92 (61.7) | 419 (61.0) |
| Coronary artery bypass graft | 57 (6.8) | 15 (10.1) | 42 (6.1) |
| Percutaneous coronary intervention | 347 (41.5) | 65 (43.6) | 282 (41.0) |
| Myocardial infarction | 101 (12.1) | 20 (13.4) | 81 (11.8) |
| Nicotine abuse | 180 (21.5) | 41 (27.5) | 139 (20.2) |
| Chronic lung disease | 185 (22.1) | 34 (22.8) | 151 (22.0) |
| Stroke | 87 (10.4) | 15 (10.1) | 72 (10.5) |
| Peripheral arterial disease | 122 (14.6) | 26 (17.4) | 96 (14.0) |
| eGFR (N = 817) | 58.0 ± 20.9 | 54.1 ± 21.3 | 58.8 ± 20.7 |
| Electrocardiography |  |  |  |
| Heart rate, bpm (N = 814) | 71 (63 - 82) | 71 (63 - 82) | 72 (63 - 82) |
| QRS complex, ms (N = 815) | 98 (90 - 114) | 104 (92 - 137) | 97 (90 - 112) |
| PQ interval, ms (N = 608) | 178 (156 - 200) | 190 (165 - 219) | 176 (155 - 197) |
| Preoperative AV block I° (N = 818) | 156 (18.7) | 42 (28.2) | 114 (16.6) |
| Preoperative LBBB (N = 818) | 69 (8.3) | 9 (6.0) | 60 (8.7) |
| Preoperative RBBB (N = 818) | 80 (9.6) | 42 (28.2) | 38 (5.5) |
| Preoperative LAHB (N = 818) | 113 (13.5) | 29 (19.5) | 84 (12.2) |
| Preoperative LPHB (N = 818) | 0 | 0 | 0 |
| Echocardiography |  |  |  |
| Ejection fraction, % (N = 780) | 60 (50 - 60) | 59 (50 - 60) | 60 (51 - 60) |
| IVSd, mm (N = 691) | 13.6 ± 3.2 | 14.4 ± 5.7 | 13.4 ± 2.2 |
| LVDd, mm (N = 660) | 45.4 ± 7.1 | 44.8 ± 7.3 | 45.5 ± 7.1 |
| Aortic valve area, cm<sup>2</sup> (N = 803) | 0.74 ± 0.20 | 0.76 ± 0.18 | 0.74 ± 0.20 |
| AV PGmean, mmHg (N = 776) | 42.0 ± 14.4 | 41.6 ± 12.9 | 42.1 ± 14.7 |
| Cardiac computed tomography |  |  |  |
| Aortic annulus perimeter, mm<br>(N = 707) | 76 (71 - 82) | 76 (72 - 81) | 77 (71 - 82) |
| MS length, mm (N = 723) | 2.4 (1.3 - 3.8) | 2.4 (1.2 - 3.4) | 2.4 (1.3 - 3.9) |
| Procedural data |  |  |  |
| Alternative access | 60 (7.2) | 9 (6.0) | 51 (7.4) |
| Balloon predilatation | 506 (60.5) | 95 (63.8) | 411 (59.8) |
| Balloon postdilatation | 217 (26.0) | 43 (28.9) | 174 (25.3) |
| Implantation depth (mean), mm | 5.1 ± 2.3 | 5.6 ± 2.5 | 5.0 ± 2.2 |
| Implantation depth (NCC), mm | 5.1 ± 2.4 | 5.6 ± 2.6 | 5.0 ± 2.4 |
| Implantation depth (LCC), mm | 5.2 ± 2.5 | 5.5 ± 2.7 | 5.1 ± 2.4 |
| Edwards Sapien 3 | 379 (45.3) | 53 (35.6) | 326 (47.5) |
| Abbott Portico | 361 (43.2) | 78 (52.3) | 283 (41.2) |
| Medtronic Evolut R/PRO | 96 (11.5) | 18 (12.1) | 78 (11.4) |
| Self-expanding valves | 457 (54.7) | 96 (64.4) | 361 (52.5) |
| Balloon-expandable valves | 379 (45.3) | 53 (35.6) | 326 (47.5) |

EuroSCORE = European System for Cardiac Operative Risk Evaluation, STS = Society of Thoracic Surgeons, NYHA = New York Heart Association, eGFR = estimated glomerular filtration rate, AV block = atrioventricular block, LBBB = left bundle branch block, RBBB = right bundle branch block, LAHB = left anterior hemiblock, LPHB = left posterior hemiblock, IVSd = interventricular septum in diastole, LVDd = left ventricular diameter in diastole, AV PGmean = pressure mean gradient over the aortic valve, MS = membranous septum, NCC = noncoronary cusp, LCC = leftcoronary cusp.
\* Values are presented as mean ± SD, median (IQR), or n (%)

### Predictors for pacemaker implantation

The first lasso regression step revealed 14 preprocedural and two periprocedural predictors (see Table 2) that correlated with PPMI. In the second lasso step, where interactions between all primarily selected variables were allowed, 16 interaction terms were added. For the prediction of PPMI, the final model (see Table 2, Model B) achieved an AUC of 0.8 with a sensitivity of 0.76 and a specificity of 0.72. We implemented the final model in an online calculator that gives individual patient and procedural risks based on our sample (https://biometrycharite.shinyapps.io/pmrisk/).

**Table 2:**
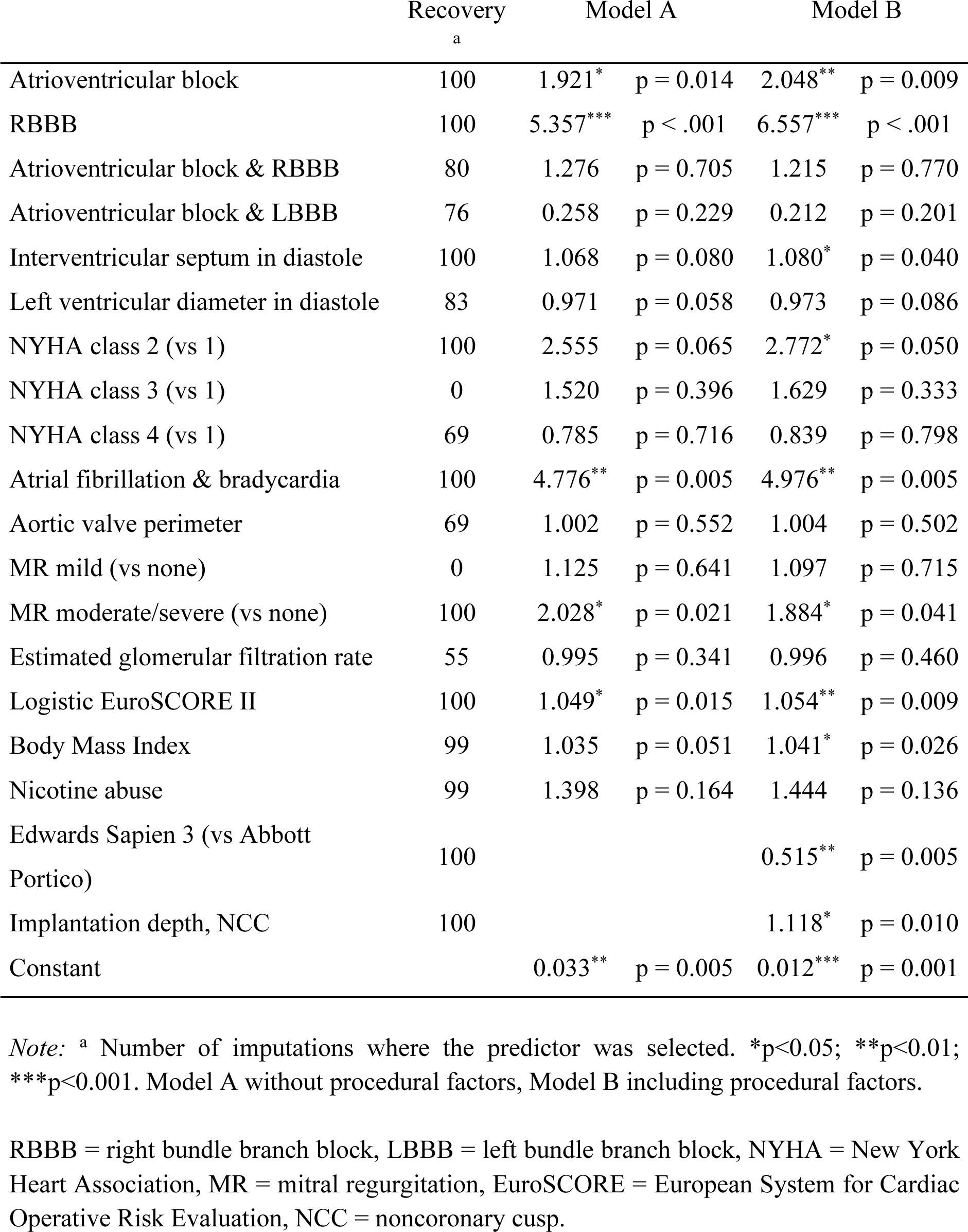
Prediction of pacemaker implantation after TAVI depending on different risk factors.

|  | Recovery<br>a | Model A | Model B |
| --- | --- | --- | --- |
| Atrioventricular block | 100 | 1.921* p = 0.014 | 2.048** p = 0.009 |
| RBBB | 100 | 5.357*** p < .001 | 6.557*** p < .001 |
| Atrioventricular block & RBBB | 80 | 1.276 p = 0.705 | 1.215 p = 0.770 |
| Atrioventricular block & LBBB | 76 | 0.258 p = 0.229 | 0.212 p = 0.201 |
| Interventricular septum in diastole | 100 | 1.068 p = 0.080 | 1.080* p = 0.040 |
| Left ventricular diameter in diastole | 83 | 0.971 p = 0.058 | 0.973 p = 0.086 |
| NYHA class 2 (vs 1) | 100 | 2.555 p = 0.065 | 2.772* p = 0.050 |
| NYHA class 3 (vs 1) | 0 | 1.520 p = 0.396 | 1.629 p = 0.333 |
| NYHA class 4 (vs 1) | 69 | 0.785 p = 0.716 | 0.839 p = 0.798 |
| Atrial fibrillation & bradycardia | 100 | 4.776** p = 0.005 | 4.976** p = 0.005 |
| Aortic valve perimeter | 69 | 1.002 p = 0.552 | 1.004 p = 0.502 |
| MR mild (vs none) | 0 | 1.125 p = 0.641 | 1.097 p = 0.715 |
| MR moderate/severe (vs none) | 100 | 2.028* p = 0.021 | 1.884* p = 0.041 |
| Estimated glomerular filtration rate | 55 | 0.995 p = 0.341 | 0.996 p = 0.460 |
| Logistic EuroSCORE II | 100 | 1.049* p = 0.015 | 1.054** p = 0.009 |
| Body Mass Index | 99 | 1.035 p = 0.051 | 1.041* p = 0.026 |
| Nicotine abuse | 99 | 1.398 p = 0.164 | 1.444 p = 0.136 |
| Edwards Sapien 3 (vs Abbott Portico) | 100 |  | 0.515** p = 0.005 |
| Implantation depth, NCC | 100 |  | 1.118* p = 0.010 |
| Constant |  | 0.033** p = 0.005 | 0.012*** p = 0.001 |

*Note:* <sup>a</sup> Number of imputations where the predictor was selected. \*p<0.05; \*\*p<0.01; \*\*\*p<0.001. Model A without procedural factors, Model B including procedural factors.
RBBB = right bundle branch block, LBBB = left bundle branch block, NYHA = New York Heart Association, MR = mitral regurgitation, EuroSCORE = European System for Cardiac Operative Risk Evaluation, NCC = noncoronary cusp.

## Discussion

Our study aimed to develop an innovative tool that assists interventional cardiologists in both selecting the appropriate THV and determining the implantation strategy based on the individual patient’s risk for PPMI. So far, most studies analyzing PPMI risk factors reported odds ratios indicating relative risks associated with certain factors. To help patients better understand their own risk – thereby facilitating shared decision-making – we designed our score to calculate the individual probability of PPMI. The risk score is easily accessible via a website. After entering the unmodifiable, preprocedural risk factors, the tool provides an initial PPMI risk assessment. In the next step, the tool calculates potential risk changes based on valve selection and intraprocedural implantation depth, allowing interventional cardiologists to adjust procedural planning according to the patient’s pacemaker risk. With this approach, we hope to contribute to a long-term reduction in the key complication of TAVI: postoperative pacemaker implantation.

The included patient group represents elderly individuals with a low-risk profile. The observed pacemaker implantation rate of 17% in our study aligns with current literature, which reports rates ranging from 3.4% to 25.9% following TAVI.^17–19^

In recent years, two risk scores for predicting PPMI after TAVI have gained attention: the risk calculation by Maeno et al. and the Emory Score by Kiani et al.^11,12^ Despite promising results of the original studies, the performance of both scores in external validation cohorts still requires improvement.^13,14^ In our study we attempted to improve certain aspects of the scores and apply a new calculating approach. The studies of Maeno et al. and Kiani et al. are both single-center studies from large American medical institutions, focusing on patients who received balloon-expandable valve types. To enhance applicability in daily clinical routine, we included both balloon-expandable and self-expanding valve types. Furthermore, the studies conducted by Maeno et al. and Kiani et al. spanned earlier periods. Considering the rapid technological advancements in recent years, our study updates the findings of Maeno et al. and Kiani et al. with more contemporary data. All our patients received treatment within a two-year timeframe, ensuring consistency in the employed valve systems and minimizing potential biases associated with postoperative conduction anomalies. Additionally, the large size of our study population and limited exclusion criteria reduced the risk of distortion. Notably, the number of patients who underwent PPMI was relatively low in both studies: 35 patients (14.6%) in Maeno et al. and 57 patients (7.3%) in Kiani et al. This smaller sample size might have made the scores highly applicable to the specific patient cohorts studied but may not fully represent the general population. With 149 patients who underwent PPMI after TAVI, we offer a larger and potentially more representative sample size.

Our statistical approach differs in two key ways. Firstly, we used multiple imputation to handle missing values. This enabled us to minimize information loss due to missing variables and to reduce bias caused by missing patterns. Secondly, we did not develop our score by using logistic regression models but rather improved its predictive power by adding lasso regression. Lasso regression is well-suited for developing risk scores because it incorporates a penalty term to prevent the model from becoming overly complex and depending too much on the training data. This approach reduces the risk of overfitting, and enhances predictive performance on new datasets.

One focus of our study was on the implantation depth as a risk factor. Maeno et al. also identified implantation depth at the NCC as a risk factor. However, Kiani et al. excluded implantation depth from their score, arguing it is measured retrospectively and therefore does not contribute to preoperative risk assessment. We attempted to address this aspect in our two-stage score by incorporating implantation depth in the second step. This approach aims to both acknowledge the point made by Kiani et al. and emphasize the significant role of implantation depth as a risk factor for PPMI.

In our risk score, we identified various variables that had a significant impact on PPMI after TAVI. AV block and right bundle branch block, well-known risk factors for PPMI after TAVI, were also strongly associated with PPMI in our analysis.^20,21^ For anatomical reasons, THV implantation primarily leads to pressure on the left Tawara branch, so pre-existing right Tawara branch blockage or pre-existing atrioventricular node conduction disturbances can lead to a correspondingly high risk for PPMI.^22^

A lower THV implantation has also been confirmed as a significant risk factor in numerous studies.^23–24^ The short distance of the conduction pathways to the distal landing zone of the THV increases their susceptibility to injury.^22,25^ The implantation depth plays a key role in avoiding PPMI as it is one of the few modifiable procedural aspects. In our study, measurement of the implantation depth between the aortic annulus and the distal end of the prosthesis at the NCC was identified as the method with the most precise differentiation of postoperative pacemaker dependency. Choosing the NCC as a reference point seems reasonable due to its anatomical proximity to the cardiac conduction system.^26^

As previously described, the implantation of a balloon-expandable valve in our study was associated with a lower risk of PPMI than the implantation of a self-expanding valve.^27,28^

The high association between logistic EuroSCORE II, elevated body mass index, and nicotine abuse with PPMI suggests that clinically multimorbid patients may have an increased risk of PPMI after TAVI. Similarly, in our study, anatomical risk factors such as the thickness of the interventricular septum and a smaller left ventricular diameter were strongly associated with PPMI. To the best of our knowledge, these risk factors have not previously been described in the context of TAVI.

In everyday clinical practice, procedural planning for TAVI is performed by the heart team based on individual patient characteristics. We aimed to reflect this aspect of individual risk stratification in our score by developing the two-stage procedure. This provides interventionalists with an overview of how certain procedural characteristics affect individual risk alongside the inherent pre-existing risk for a pacemaker.

## Study Limitations

The results of this retrospective cohort study serve as empirical evidence to strengthen hypotheses, but they cannot prove a causal relationship. The THV choice was the responsibility of the interventional cardiologist. Individual patient risk profiles, anatomical characteristics, and the current knowledge of postoperative conduction anomaly risk factors likely influenced THV selection and may have impacted the pacemaker rate after TAVI. Notably, we lacked information regarding the pacing rate after TAVI. Besides the identified risk factors, there may be unknown factors that have influenced the pacemaker rate after TAVI.

## Conclusion

Our risk score fills the existing data gap regarding newer prosthesis generations and introduces a novel approach to risk score calculation. The two-step nature of our score allows interventional cardiologists, to initially calculate the patient’s individual pacemaker risk before TAVI based on unmodifiable clinical risk factors. Building upon this outcome, our risk score enables them to assess how the pacemaker risk of their patient can effectively be influenced by specific valve types or implantation techniques. This two-step risk score may provide a novel and valuable tool for daily clinical practice.

## Data Availability

All data utilized in this study can be requested and viewed by the authors.

## Acknowledgments

1. D. L. with H. D. had the idea for this study. Data acquisition was performed by a group of seven investigators (A. J., J. L, B. J., L. M., M. H., P. H.). The statistical analysis was performed by D. S. The manuscript was mainly authored by A. J. and J. L., all authors discussed the results and contributed to the final manuscript.

## Non-standard Abbreviations and Acronyms

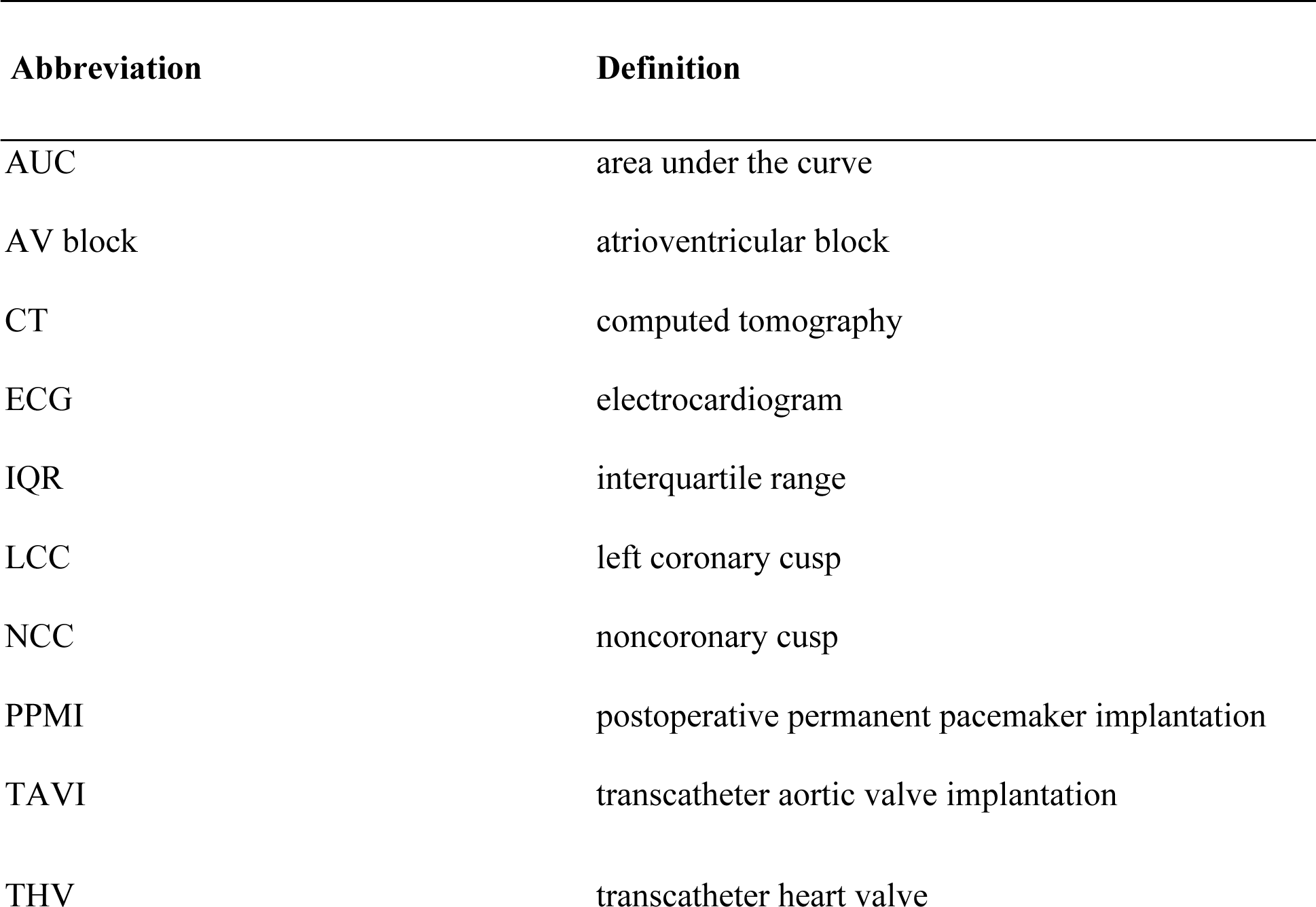

| Abbreviation | Definition |
| --- | --- |
| AUC | area under the curve |
| AV block | atrioventricular block |
| CT | computed tomography |
| ECG | electrocardiogram |
| IQR | interquartile range |
| LCC | left coronary cusp |
| NCC | noncoronary cusp |
| PPMI | postoperative permanent pacemaker implantation |
| TAVI | transcatheter aortic valve implantation |
| THV | transcatheter heart valve |

